# Early experience with modified dose nirmatrelvir/ritonavir in dialysis patients with coronavirus disease-2019

**DOI:** 10.1101/2022.05.18.22275234

**Authors:** Pierre Antoine Brown, Michaeline McGuinty, Christos Argyropoulos, Edward G Clark, David Colantonio, Pierre Giguere, Swapnil Hiremath

## Abstract

**Introduction:** Nirmatrelvir/Ritonavir was approved for use in high risk outpatients with coronavirus disease (COVID-19). However, patients with severe chronic kidney disease, including patients on dialysis, were excluded from the phase 3 trial, and currently the drug is not recommended below a glomerular filtration rate of 30 ml/min/1.73m^2^. Based on available pharmacological data and principles, we developed a modified dose which was lower, and administered at longer intervals.We administered nirmatrelvir/ritonavir as 300/100 mg on day one, followed by 150/100 mg daily from day two to day five. In this case series, we report the initial experience with this modified dose regimen.

**Methods:** This is a retrospective chart review, conducted after obtaining institutional board approval. Demographic and outcome data was abstracted from the electronic medical record for dialysis patients who developed COVID-19 during the period of study and received nirmatrelvir/ritonavir. The principal outcomes we describe are symptom resolution, and safety data with the modified dose regimen in the dialysis patients.

**Results:** 19 patients developed COVID-19 during the period of study of whom 15 received nirmatrelvir/ritonavir. 47% of them were female and 67% had diabetes. Most patients had received three doses of the vaccine (80%) while 13% were unvaccinated. Potential drug interactions concerns were common (median 2 drugs per patient) with amlodipine and atorvastatin being the commonest drugs requiring dose modification. Nirmatrelvir/ritonavir use was associated with symptom resolution in all patients, and was well tolerated. One patient had a rebound of symptoms, which improved in 2 more days. There were no COVID-19 related hospitalizations or deaths in any of the patients.

**Conclusion:** In this case series of 15 hemodialysis patients with COVID-19, a modified dose of nirmatrelvir/ritonavir use, with pharmacist support for drug interaction management, was associated with symptom resolution, and was well tolerated with no serious adverse effects.

## Introduction

As of May 15, 2022, there have been 6.27 million deaths globally from Coronavirus disease (COVID-19) due to the Serious Acute Respiratory Syndrome Coronavirus-2 (SARS-CoV-2),with estimates of excess mortality being about 3 times higher at 18.2 million (1,2). Additionally, there are long term consequences of COVID-19, including post-critical illness syndromes, auto-immune diseases, cardiovascular disease, and other physical and neurocognitive sequelae (3–5). COVID-19 infections and their consequences will occur over the next few years and will have a major impact on morbidity, life expectancy and healthcare utilization. The burden of morbidity and mortality of COVID-19 is higher in immunocompromised people including those with advanced CKD (stages 4 and 5) and those with end stage kidney disease (ESKD), receiving dialysis. Hemodialysis patients cannot self-isolate, usually receiving their life-sustaining treatment three times a week in a congregate setting. Presently, about 3 million people receive dialysis worldwide, 23,708 of them in Canada (6,7). In the first year after the declaration of the pandemic, in the United States alone, the year-over-year decline in the dialysis census was 1.6%, representing a deficit of 3.8%, relative to the forecasted increasing trend (8).

The vaccines for COVID-19 are remarkably effective, however their efficacy is lower in the dialysis population. The pooled estimate of early antibody formation in dialysis patients was 89% (95% confidence interval [CI] 85 to 91%) (9) relative to healthy controls, conferring incomplete protection which wanes over time (10–12). For evolving variants (such as Omicron) which require higher antibody titres for viral neutralization, a corollary is that vaccines alone will not be sufficient for protection against infection and severe disease in dialysis patients (13). Several therapeutic options have been investigated, and some have been approved for early treatment of COVID-19, notably remdesivir (Velkury, Gilead Inc) and nirmatrelvir/ritonavir (Paxlovid, Pfizer Inc.). However, patients with low GFR have been excluded from all clinical trials of nirmatrelvir/ritonavir, and theoretical concerns about drug dosing and safety have led to glomerular filtration rate (GFR) < 30 ml/min/1.73m^2^ being given a ‘do not recommend’ label in the product monograph

Patients with chronic kidney disease (CKD), especially those with GFR < 30, dialysis and transplant recipients are frequently excluded from clinical trials, in particular those evaluating investigational drugs. Proposed explanations for this include that the high comorbidity burden in these populations may introduce confounding unrelated outcomes. Additionally, decreased renal clearance for newer drugs makes dosing decisions difficult for a phase 3 trial. The consequence is that CKD patients are excluded initially at time of drug approval, and therapeutic nihilism occurs, where these patients are denied effective therapies until data emerges supporting safety and efficacy, often many years later (14,15). This phenomenon has been termed ‘renalism’, and unfortunately continues with COVID-19. A rapid review of trial registries for COVID-19 reported that 218 of 484 trials (45%) had an exclusion based on CKD status which was often poorly defined (such as ‘kidney dysfunction’ without a GFR cutoff) (16).

On the basis of available pharmacological data for nirmatrelvir/ritonavir, an adjusted therapeutic regimen with a lower dose and longer interval between doses can achieve required drug concentration in the serum (17). The available data also does not suggest dose-dependent toxicity. Given the risk of severe disease in the hemodialysis population and the lack of alternative treatment options for the current, and projected future circulating variant in Ontario, we implemented a modified nirmatrelvir/ritonavir dosing protocol at our institution. In this report, we provide the preliminary safety and efficacy data in this cohort of dialysis patients with COVID-19 receiving lower dose nirmatrelvir/ritonavir.

## Methods

### Setting

We conducted a retrospective chart review of hemodialysis patients at our institution who received nirmatrelvir/ritonavir. The protocol was implemented on April 5th, and we report on the first 15 patients who received this drug for COVID.

### Population

Patients were considered eligible for outpatient COVID therapeutics based on the Ontario Science Table guidance. Briefly, outpatient therapy for COVID is considered for unvaccinated patients and vaccinated patients who are at higher risk for adverse outcomes due to underlying risk factors including immunosuppressed status, comorbid conditions, and age. Being on dialysis is considered one of these risk factors. Patients have to be symptomatic, but not hospitalized, to be considered for these outpatient therapies.

### Procedures

Until April 11, 2022, eligibility for outpatient COVID therapy required adjudication and approval by an institutional committee which included an infectious diseases physician (MM). This requirement was removed on April 12, 2022. Two nephrologists (PAB, SH) were notified of all dialysis patients with a COVID diagnosis. At our institution, All hemodialysis patients with COVID are cohorted at an evening shift at a single unit.. After reviewing the history for eligibility of receiving outpatient therapy (presence of symptoms and the ability to start therapy within specified duration), the medication list was reviewed for relevant drug-drug interactions by expert pharmacists. Patients using contraindicated drugs such as amiodarone or CYP450 inducers (e.g. anticonvulsants) were offered alternate therapy options (e.g. remdesevir). The potential drug-drug interactions (e.g. amlodipine, atorvastatin) were managed through dose reduction or interruption of the drug based on clinical judgment. The details of the process are shown in figure 1.

**Figure 1:**
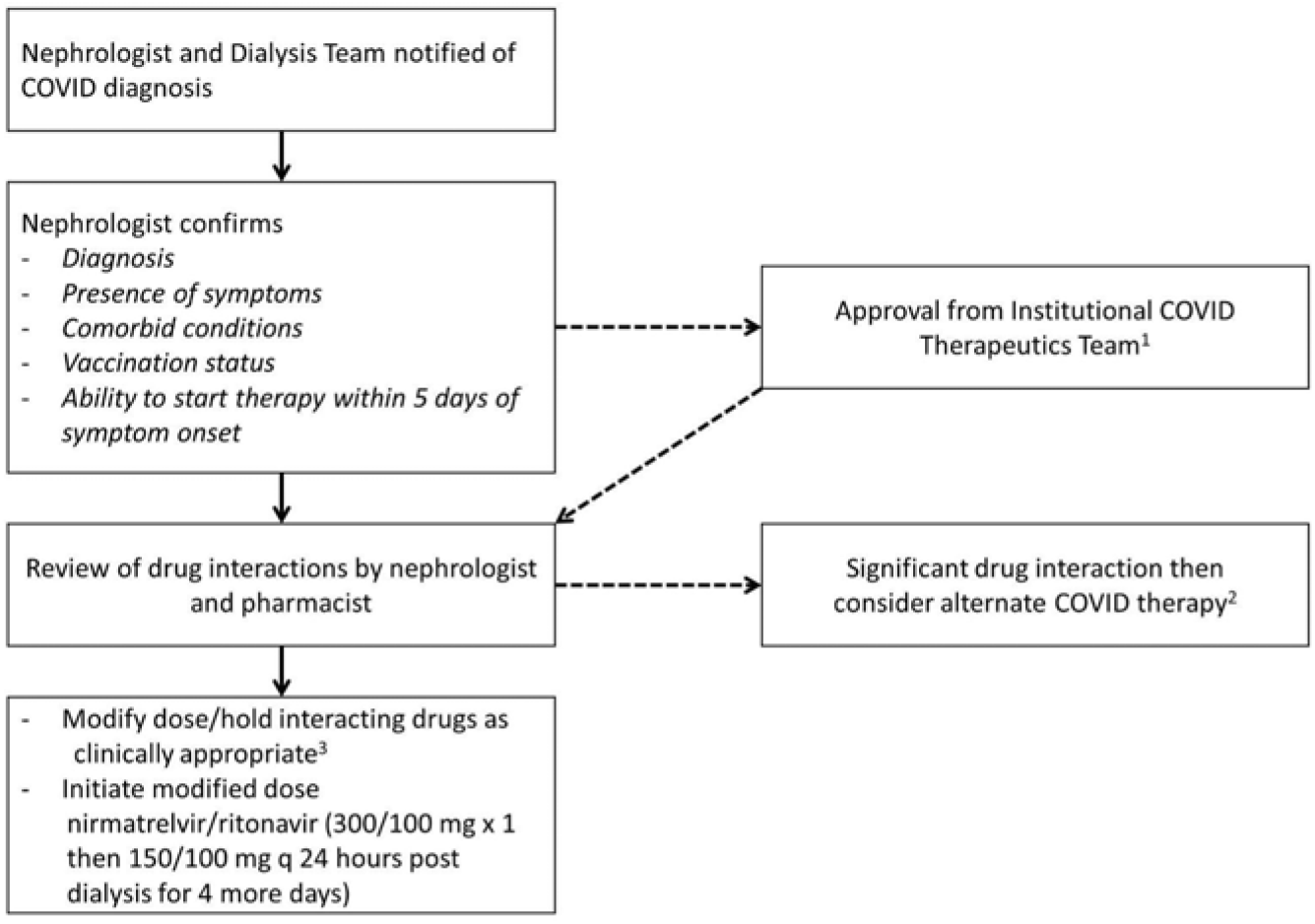
Procedures for Prescribing Nirmatrelvir/Ritonavir in Dialysis.

### Rationale for modified dose

A single dose of nirmatrelvir 100 mg provided adequate concentration of drug to inhibit Mpro enzymatic activity at 24 hours, based on study C4671005 in non-dialysis CKD patients with GFR < 30 (17). Similar concentrations would be seen in dialysis, and it is expected that there will be some dialytic clearance of nirmatrelvir, given its molecular size, 70% protein binding and volume of distribution. The safety profile of nirmatrelvir is quite favorable, with few serious adverse effects, and the animal data does not indicate a dose dependent toxicity. Nirmatrelvir is currently formulated as 150 mg and dosed at 300 mg with 100 mg ritonavir twice a day for patients with normal kidney function, and at 150 mgs with 100 mg ritonavir twice a day in those with GFR 30-60. Hence a dose of 300 mg (with 100 mg ritonavir) followed by 150 mg daily, administered after dialysis on dialysis days would be effective for enzyme inhibition. The data from study C4671005 is after a single dose, and it is expected that the drug will accumulate with repeat doses, however the duration of therapy is 5 days which would limit this and any cumulative toxicity.

### Analysis

Institutional review board approval was obtained before collecting these data for this report from the Ottawa Health Sciences Research Ethics Board. Data was extracted from the electronic medical record (EPIC Systems, Verona, Wisconsin) for these patients. We report summary baseline characteristics including demographics, symptoms, and safety and efficacy data for the first 15 patients treated at our institution.

## Results

Over the period of study, 19 hemodialysis patients developed COVID-19, of whom two were asymptomatic, and two were using drugs that are contraindicated, hence 15 patients were started on nirmatrelvir/ritonavir (figure 2).

**Figure 2:**
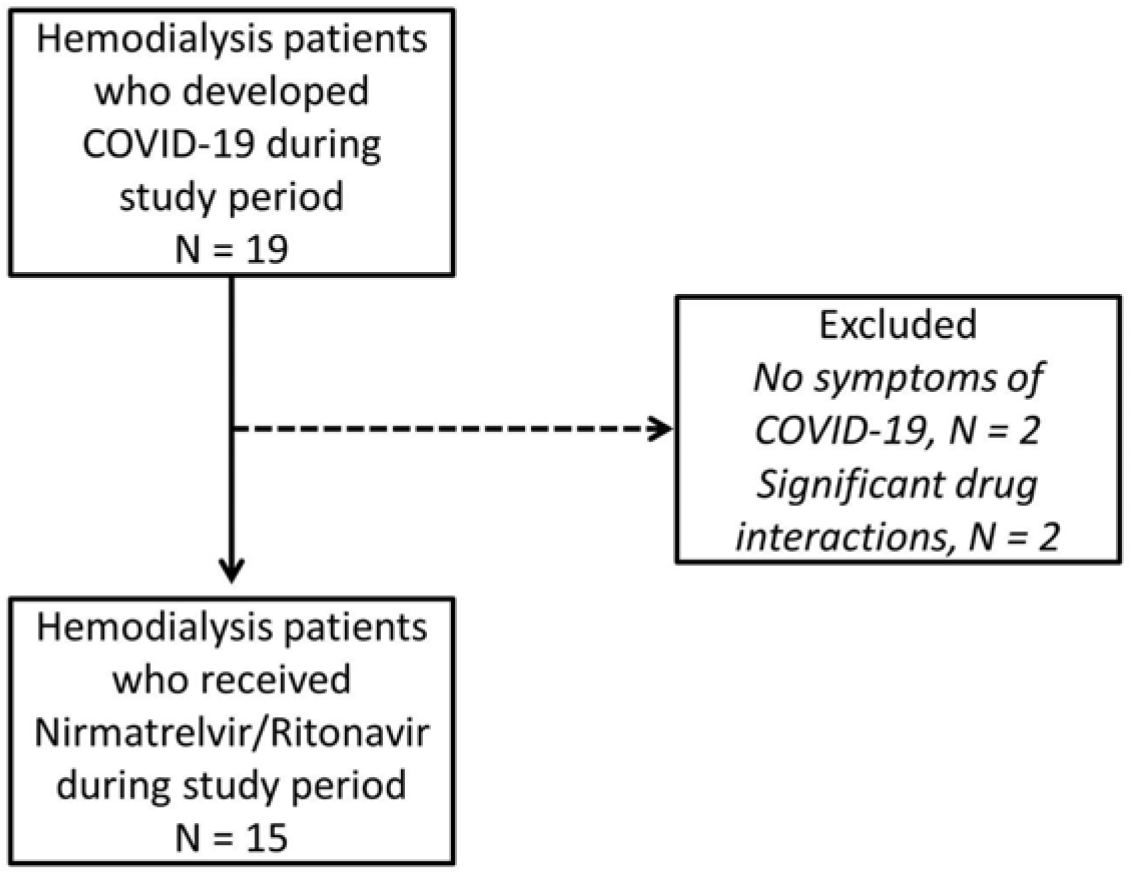
Flow of patients included in this study.

The baseline characteristics for these 15 patients are provided in table 1. Two-thirds of them had diabetes, and 3 had a previous failed transplant and were on prednisone. One patient had received rituximab 4 months prior and was also receiving prednisone. Two patients were unvaccinated, 12 patients had received 3 doses and only one patient had received 4 doses of the vaccine. Cough and runny nose were the commonest symptoms. The median time from onset of symptoms to initiation of therapy was 2 days.

**Table 1:** Baseline characteristics of patients who received nirmatrelvir/ritonavir.

| Characteristic | Value |
| --- | --- |
| Total number | 15 |
| Female sex | 7 (47%) |
| Age | 60.2 ± 15.2, range 34 to 84 |
| Diabetes | 10 (67%) |
| Other comorbidities |  |
| *Hypertension* | 13 (87%) |
| *Coronary Artery Disease* | 4 (30%) |
| *Peripheral Arterial Disease* | 3 (20%) |
| *Previous failed Transplant* | 3 (20%) |
| Immunosuppressive agents | 4 (3 prednisone; 1 rituximab and prednisone) |
| Vaccine doses |  |
| *Unvaccinated* | 2 (13%) |
| *3 doses* | 12 (80%) |
| *4 doses* | 1 (7%) |
| Symptoms |  |
| *Cough* | 11 |
| *Nasal congestion/runny nose* | 8 |
| *Fatigue/malaise* | 6 |
| *Fever* | 4 |
| *Diarrhea* | 2 |
| *Others\** | 4 |
| Days from symptom onset to initiation of therapy | Median 2 days (range 1 to 4) |
| Number of drugs with interactions | Median 2 (range 0 to 4) |

\* One each with shortness of breath (not hypoxemic), hemoptysis, headache and sore throat

### Drug-Drug Interactions

Most patients did have drugs that would need adjustment due to interactions with ritonavir. The median number of drugs requiring adjustment was 2, and the commonest drugs requiring adjustment were atorvastatin (8) and amlodipine (6). No adverse effects related to drug dose adjustments were noted.

### Outcomes

All patients reported an improvement and resolution of symptoms within the 5 days of therapy. In one patient, an initial resolution of symptoms was followed by a ‘rebound’ of symptoms at day 7, which resolved again 10 days after onset of symptoms.

### Safety

Nirmatrelvir/ritonavir was well tolerated and none of the patients had to withdraw the drug for intolerance. One patient noted a headache and two others mentioned a change in taste. One patient died 10 days after initial diagnosis, and 2 days after completing nirmatrelvir/ritonavir. Their symptoms had resolved after starting the drug. They had a cardiac arrest just at the end of the long interdialytic interval, and a serum potassium was noted to be 7.3 at the time, suggesting this was unrelated to COVID-19 or the drug therapy. None of the other patients required hospitalization.

## Discussion

In this case series we describe the initial experience with the use of a modified dose of nirmatrelvir/ritonavir in dialysis patients. The drugs were well tolerated, all patients had resolution of symptoms, and none required COVID-19 related hospitalization. Most patients did require adjustment of drugs to pre-empt the potential effects of drug interactions with ritonavir. This case series supports the use of modified dose nirmatrelvir/ritonavir in dialysis patients as being well tolerated with efficacy in the form of symptom relief.

This is the first description of nirmatrelvir/ritonavir in dialysis patients. Currently, the drug monograph suggests that these drugs are ‘not recommended’ with serious renal impairment, given concerns of accumulation of drugs and lack of data. Based on pharmacological principles, we decided to use a lower dose at a longer interval in dialysis patients in this case series. In this ongoing seventh wave in Ontario, the BA2 variant of Omicron is predominant, and the therapeutic options were severely limited as a consequence. Based on *in vitro* neutralization assays, this variant is not neutralized by sotrovimab or prior monoclonal antibodies. The other oral antiviral agent, molnupiravir, is not available yet in Canada. Lastly, remdesivir also has been reported to reduce hospitalization in outpatients at high risk, however there were no dialysis patients included in the phase 3 trial, either (18). Indeed, in the two patients in whom we felt the drug interactions were severe enough and we did not use nirmatrelvir/ritonavir, we did use remdesivir.

Rebound of symptoms was noted in one of the 15 patients (5%), however this improved spontaneously after two more days.In the phase 3 clinical trial, some patients (range 1-2%) had one or more positive SARS-CoV-2 PCR tests after testing negative, or an increase in the amount of SARS-CoV-2 detected by PCR, after completing their treatment course (19). Our experience seems higher than that, though an imprecise estimate given small numbers, and of uncertain significance. Some have advocated either a longer course, or a repeat course of nirmatrelvir/ritonavir if rebound phenomenon develops, but robust evidence for the explanation and treatment is lacking.

This study does have limitations. Most importantly, this is not a randomized controlled trial, and we do not have controls for comparison. Comparing these data with historical controls would be biased, given changing vaccination status, variants, and therapeutic options over time. A comparison with patients who did not receive nirmatrelvir/ritonavir also would be biased, as these patients would be importantly different from the treated group.. Lastly, the purpose of this case series is not to demonstrate the efficacy of nirmatrelvir/ritonavir, rather to report that a modified dose is well tolerated in this population.

The importance of our experience is to allow faster implementation of modified dose protocol of nirmatrelvir/ritonavir for dialysis patients and shorten the period of therapeutic nihilism. Though monoclonal antibodies are easier and considered safer to use in the dialysis population, since there is no need for dose adjustment, their efficacy also changes as newer and resistant variants arise with mutations in the receptor binding domain (RBD) of the spike protein. This is less likely to happen with antiviral drugs, and the experience we describe will be useful for management of dialysis patients in the next few waves.

We do not have robust evidence in the form of a randomized controlled trial. It should be noted that the phase 3 trial which led to the drug approval included 2246 patients, and to do such a large trial specifically in dialysis patients may not be feasible (20). Waiting for trial evidence in this specific population would also mean a delay in the availability of an effective therapy in a vulnerable population which is at higher risk for contracting COVID-19 as well as experiencing higher subsequent morbidity and mortality. Phase 3 trials should not reflexively exclude patients with severe kidney disease, and should incorporate modified doses based on pharmacological rationale and/or pharmacokinetic studies. Until that becomes a reality, the renal community needs to embrace quicker implementation of modified dose protocols based on pharmacological principles.

## Conclusion

In this case series of 15 hemodialysis patients with COVID-19, a modified dose regimen of nirmatrelvir/ritonavir use was associated with symptom resolution, and was well tolerated with no serious adverse effects

## Data Availability

All data produced in the present study are available upon reasonable request to the authors, and after approval from the Ottawa Health Sciences Research Ethics Board

## Acknowledgments

This study had no direct funding. SH, EGC, PAB receive research salary support from the Department of Medicine, University of Ottawa.

## Notes

### Competing Interest Statement

The authors have declared no competing interest.

### Funding Statement

The study did not receive any direct funding. SH, PAB and EGC receive research salary support from the Department of Medicine, University of Ottawa.

### Author Declarations

The Institutional review board of the Ottawa Hospital, the Ottawa Health Sciences Research Ethics Board gave ethical approval for this work.

